# Effects of Chronic Pain Diagnoses on the Antidepressant Efficacy of Transcranial Magnetic Stimulation

**DOI:** 10.1101/2023.06.27.23291964

**Authors:** H. Totonchi Afshar, J. N. Fishbein, E. J. Martinez, G. M. Chu, M. A. Shenasa, D. Ramanathan, M. S. Herbert

## Abstract

**Background:** Major depressive disorder (MDD) and chronic pain are highly comorbid and bidirectionally related, such that MDD typically interferes with chronic pain treatment and vice versa. Repetitive transcranial magnetic stimulation (rTMS) over the dorsolateral prefrontal cortex (DLPFC) is effective in treating MDD, but additional research is needed to determine if chronic pain interferes with rTMS for MDD.

**Method:** Participants were 124 veterans (M_age_=49.1, SD=13.8) scheduled for 30 sessions of rTMS across six weeks at the Veterans Affairs San Diego Healthcare System. Depression severity was monitored weekly using the Patient Health Questionnaire-9. Having any pain diagnosis, low back pain, or migraine/headache were assessed by chart review. Latent basis models were used to estimate change and change-by-pain diagnosis in depression scores during rTMS treatment.

**Results:** A total of 92 participants (74%) had a documented pain diagnosis, 58 (47%) had low back pain, and 32 (26%) had migraine/headache. Depression scores initially decreased (linear slope estimate=-2.04, SE=0.26, *p*<.0001), but the rate of decrease slowed over time (quadratic slope estimate=0.18, SE=0.04, *p*<.001). Having any pain diagnosis, low back pain, or migraine/headache did not significantly differentiate overall amount of change. However, individuals with headache/migraine showed greater initial improvement but then an even faster slowing in rate of decrease than those without headache/migraine.

**Conclusions:** Having any pain diagnosis, low back pain, or headache/migraine did not significantly interfere with improvement in depression; however, headache/migraine affected the timing of change. These data contribute to the ongoing support of rTMS as a viable treatment option for comorbid populations.

## Introduction

Major depressive disorder (MDD) and chronic pain are two of the most prevalent and disabling health conditions worldwide. MDD is a mood disorder that affects more than 300 million people globally (World Health Organization, 2017), while chronic pain, defined as pain that lasts for more than three months, affects an estimated 20% of adults (Nahin, 2015). Both conditions can have a profound impact on quality of life, social functioning, and economic productivity (Gureje et al., 1998; Kawai et al., 2017; Richards, 2011). There has been a longstanding interest in the relationship between MDD and chronic pain (Dworkin & Gitlin, 1991; Haythornthwaite et al., 1991) and there is a high degree of comorbidity between the two conditions: up to 85% of patients with chronic pain also experience symptoms of depression and approximately 65% of patients with MDD report pain as a common symptom (Bair et al., 2003, 2008). Further, the two conditions are bidirectionally related, as depression can exacerbate chronic pain severity and vice versa (Arnow et al., 2006; Fishbain et al., 1997; Kroenke et al., 2011).

Although the exact mechanisms underlying the high overlap between MDD and chronic pain are not fully understood, both MDD and chronic pain are associated with alterations in cortico-limbic neural areas (e.g., anterior cingulate cortex), neurotransmitter concentrations (e.g., serotonin, norepinephrine), and cognitive factors (e.g., decreased self-efficacy)(Surah et al., 2014). While this pattern of neurological overlap implies that treatments for one condition may also be effective for the other, the majority of research shows that MDD in fact interferes with pain treatment (Greco et al., 2004; Leuchter et al., 2010; Teh et al., 2010). Although the effect of chronic pain on MDD treatment have been relatively less studied, studies similarly suggest that the presence of chronic pain limits the effectiveness of MDD treatment (Ang et al., 2009). This includes traditional pharmacological interventions for MDD such as selective serotonin reuptake inhibitors, with studies suggesting the presence of chronic pain predicts poorer antidepressant medication response (Bair et al., 2003, 2004; Gerrits et al., 2012).

Transcranial magnetic stimulation (TMS) is a non-invasive technique that uses magnetic fields to stimulate nerve cells. TMS over the dorsolateral prefrontal cortex (DLPFC), a neural region implicated in cognitive processing and top-down control of emotional responding, has been shown to be effective in treating MDD, with studies reporting response rates of up to 50-60% (George et al., 2013; O’Reardon et al., 2007). TMS applied over the DLPFC or motor cortex is also effective in treating chronic pain (Yang & Chang, 2020). Thus, TMS may be a promising non-pharmacological treatment option for patients with comorbid MDD and chronic pain. While some studies suggest that TMS may be effective for reducing both depression symptoms and pain in patients with comorbid MDD and chronic pain (Leung et al., 2018; Short et al., 2011), other studies have reported mixed results (Hsu et al., 2018) or suggest that TMS may be less effective for patients with comorbid pain (Corlier et al., 2023).

Given the prevalence of comorbid MDD and chronic pain, and in light of their overlapping pathophysiology, more research is needed to determine the efficacy of TMS for patients with comorbid MDD and chronic pain and to identify which patients are most likely to benefit from this treatment approach. The primary objectives of this study were thus to: (1) evaluate the effect of pain comorbidity on rTMS treatment response for MDD; and (2) evaluate differences in rTMS treatment outcome based on the type of chronic pain. Specifically, we were interested in low back pain and headache given its high occurrence in military Veterans (Altalib et al., 2017; Radomski et al., 2020).

## Methods

### Participants

Data for the present study was collected by chart review and included 124 Veterans (M_age_=49.14, SD=13.83; female gender: n=27 [22%]) that received rTMS at the VA San Diego Healthcare System (VASDHS) Neuromodulation Clinic between February 2018 and May 2022. All Veterans were initially referred to the Neuromodulation Clinic by their primary psychiatrist. Referral eligibility was generally a failure of at least one antidepressant medication. Exclusion criteria for rTMS were as follows: no history of seizures/seizure disorder; no metallic/electrical objects implanted above the head/neck; lack of imminent suicidality and lack of recent substance dependence/misuse (within the last 2 months). Veterans were permitted to continue prescribed medications during rTMS treatment but were asked to abstain from changing medications. After eligibility was verified, Veterans were scheduled for five rTMS sessions per week for a total of 30 treatments. This retrospective analysis was approved as an IRB exemption by the VASDHS IRB.

### Assessments

#### Self-Report of Depression

As part of routine clinical practice within the VASDHS Neuromodulation Clinic, Veterans were administered the Patient Health Questionnaire-9 (PHQ-9) (Kroenke et al., 2001) at the start of rTMS and weekly thereafter during scheduled treatments to monitor depression symptoms. Since participants could receive up to six weeks of rTMS (see *Stimulation Protocol* below), each participant provided a pre-treatment score and up to six follow-up scores on these measures. The PHQ-9 is a reliable and valid measure of depression severity that contains nine items answered on a 0-point (not at all) to 3-point (nearly every day scale) scale with a range of 0 – 27.

#### Chart Review of Pain Diagnoses

We included all pain diagnoses that were listed in the Veteran’s medical chart as long as the diagnosis was made prior to the first rTMS treatment (see Supplemental Table 1 for a list of pain diagnoses). For the purposes of analysis, past yes/no pain diagnoses were coded as 0 = no past diagnosis and 1 = documented past pain diagnosis. Similarly, we coded whether participants had a diagnosis of low back pain or headache/migraine (0 = no, 1 = yes).

### Stimulation Protocol

Patient received either a 10 Hz or intermittent Theta Burst Stimulation protocol (iTBS). Both 10 Hz rTMS and iTBS entailed five days/week of treatment for up to 30 days of treatment. Both protocols entailed stimulation delivered to the left DLPFC typically at 120% of the motor threshold. The 10 Hz protocol used in the clinic was specified as 10 pulses per second for four seconds (40 total pulses/train), followed by a 12-26-second intertrain interval. We delivered 75 trains, lasting from between 19-37.5 minutes per day of stimulation for a total of 3000 pulses per session (George et al., 2010). The iTBS protocol used in the clinic was specified as a 50 Hz burst of three pulses, with bursts timed to occur every 200ms for two seconds, (40 pulses/train), followed by an eight-second intertrain interval. 20 such trains were delivered for a total of 600 pulses (Huang et al., 2005), and this stimulation protocol lasted approximately three minutes. We included four individuals in the iTBS group who received bilateral treatments (two with iTBS on the right, and two with 1 Hz treatment on the right). Individuals who were receiving other stimulation protocols (exclusively right-sided treatment or 1 Hz treatment, for example) were excluded from further analysis. There was no difference in the antidepressant response for the two stimulation protocols and thus for this analysis we combined protocols (Shenasa et al., 2023).

### Statistical Analyses

We assessed the association among age, gender, any pain diagnosis, low back pain, headache/migraine with baseline PHQ-9 scores using chi-square, independent samples t-tests, and linear regression as indicated. We used latent basis models to estimate the trajectory, overall amount of change, and overall change-by-pain diagnosis interaction in PHQ-9 scores during the intervention. Latent basis models provide estimates at each timepoint of the total change percentage from the first to last timepoint. We estimated one unmoderated latent basis model and separate moderated latent basis models each with one past pain diagnosis entered as a predictor of the intercept and slope. Then, we used these percentage estimates to ascertain whether a linear, quadratic, or some other function might capture the change over time in PHQ-9 scores. Both linear and quadratic functions appeared plausible (see *Results* below); we thus estimated linear and quadratic latent curve models and found that the quadratic models provided superior fit (*p* < .001). We fit moderated quadratic models (one model per moderator) to examine whether linear and quadratic change estimates differed by pain diagnosis. Missing data rates on the PHQ-9 were as follows: 6 missing observations (5% of total *N* = 124 sample) at baseline, 17 (14%) at week 1, 18 (15%) at week 2, 17 (14%) at week 3, 28 (23%) at week 4, 36 (29%) at week 5, and 60 (48%) at week 6. The latent basis and curve models were estimated using full information maximum likelihood estimation in the R *lavaan* package (Rosseel, 2012) version 0.12.

## Results

### Pain Diagnoses

Of the 124 participants, 92 (74%) had some documented pain diagnosis. Fifty-eight (47%) had a documented diagnosis of low back pain and 32 (26%) had documented headache/migraine. Twenty-four (19%) of participants had just one documented pain diagnosis, whereas 25 (20%) had two diagnoses, 21 (17%) had three diagnoses, and 22 (18%) had four diagnoses.

Gender, having any pain diagnosis, low back pain, and headache/migraine was not associated with baseline depression severity. Age was marginally inversely associated with baseline depression severity (*r* = .16, *p* = .09). Total number of pain diagnoses was marginally associated with greater baseline depression severity (*b* = 0.70, SE = 0.36, *p* = .06). On average, participants completed the majority of scheduled TMS sessions (M=27.08, SD=5.40), and there were no differences in number of completed sessions among participants with and without a pain diagnosis.

### Total Amount of Change on Depression Symptoms

The unmoderated latent change model indicated that participants achieved 45.2% of the total amount of improvement on depression scores by the first follow-up timepoint, 71.0% by the second follow-up, 77.9% by the third follow-up, and 87.6% by the fourth follow-up. The moderated latent basis models indicated that the overall amount of change did not differ among individuals with any past pain diagnosis (estimate = 1.00, SE = 1.26, *p* = .42), with history of low back pain (estimate = 1.78, SE = 1.11, *p* = .11), or with history of headache/migraine (estimate = -1.40, SE = 1.29, *p* = .28), relative to those without the respective pain condition.

### Timing of Depression Symptom Changes

Consistent with findings from the latent basis models, an unmoderated quadratic model of PHQ-9 scores indicated that participants’ scores initially decreased (linear slope estimate = -2.04, SE = 0.26, *p* < .001), but that the rate of decrease slowed over time (quadratic slope estimate = 0.18, SE = 0.04, *p* < .001). Individuals with a past diagnosis of headache/migraine showed greater initial improvement (linear slope-migraine interaction estimate = -1.43, SE = 0.56, *p* = .01) but then an even faster slowing in rate of decrease (quadratic slope-migraine interaction estimate = 0.22, SE = 0.78, *p* < .01) than those without headache/migraine. Other interactions between slope and past pain diagnoses were nonsignificant.

## Discussion

Given the high comorbidity and bidirectional influence of chronic pain and MDD, this study sought to examine pain as a moderator of rTMS-related improvement in depression symptomology among Veterans receiving a standard rTMS clinical protocol. The majority of Veterans had a pain diagnosis, about half had a low back pain diagnosis, and 26% had a headache/migraine diagnosis. Significant improvement in depression severity was observed across participants. Having any pain diagnosis or low back pain did not significantly differentiate response rates relative to Veterans without pain or low back pain, respectively. Among Veterans with headache/migraine, an initial steeper decrease in depression severity was observed relative to Veterans without headache/migraine, but this rate subsequently slowed such that both groups were similar by the end of treatment. Taken together, our results suggest that the presence of any pain, low back pain, or headache/migraine may not impact the reduction of depression symptoms associated with rTMS.

Our findings are consistent with previous literature showing rTMS over the DLPFC is effective in reducing depression severity in individuals with chronic pain (Freigang et al., 2021; Kumar et al., 2018; Short et al., 2011). This includes findings by Corlier et al. (2021), who showed that rTMS over the DLPFC improved both depression and pain severity ratings among 162 outpatients with MDD and ongoing pain; however, response rates in this study, defined as greater or equal to 50% decrease on the Inventory of Depressive Symptomatology Self Report version, were lowest among patients reporting severe pain severity. Corlier et al. did not examine pain diagnosis as a moderator. When considered with the results of the present study, the severity of current pain (which was not available in the current retrospective study dataset) may be a more important predictor of rTMS treatment response than the diagnosis of a pain condition. Future research is encouraged to assess both pain diagnosis and intensity of current pain as a moderator of rTMS treatment outcomes.

While the rate of change in depression severity was not significantly different among Veterans with any pain diagnosis or low back pain, Veterans with a headache/migraine diagnosis exhibited a more rapid improvement in depression severity than Veterans without headache/migraine. This was an unexpected finding that is in need of replication. rTMS treatment protocols designed to treat headache/migraine tend to be shorter than rTMS treatment protocols for depression (Leung et al., 2020) and, although the motor cortex is the preferred stimulation area in headache/migraine, a recent systematic review and meta-analysis showed the rTMS over the DLPFC for migraine was associated with reduced medication use and functional disability compared to sham stimulation (Kumar et al., 2018). In the current study, it is plausible that Veterans with headache/migraine experience a synergistic effect of rTMS that led to a more rapid improvement in depression.

The strengths and limitations of the current study are worth considering. The study sample was a relatively large group of representative Veterans. Veterans tend to have worse physical and mental health than civilians (Agha et al., 2000) and poorer treatment response to rTMS for depression (Yesavage et al., 2018), and therefore in need of additional research to elucidate mechanisms that may interfere with treatment. Results of the present study suggest that rTMS is effective for Veterans receiving rTMS for depression regardless of pain diagnosis. The study also took place within a naturalistic clinical setting and therefore may better generalize to routine clinical practice. Regarding limitations, we were unable to assess current pain severity or determine the impact of rTMS on pain. Additionally, we were unable to examine the duration of rTMS treatment effects and if this differs by pain status.

MDD and chronic pain are highly comorbid and the presence of one condition often interferes with the treatment of the other. Notably, about three-quarters of individuals seeking rTMS in the current study sample had some pain diagnosis. Here, we show that having any pain diagnosis, low back pain, or headache/migraine did not significantly influence the reduction of depression symptoms associated with rTMS over the DLPFC. These data contribute to the ongoing support of rTMS as a viable treatment option for comorbid populations, though future research is needed to better understand the extent to which the intensity of current pain affects treatment response as well as the duration of rTMS treatment effects.

## Data Availability

All data produced in the present study are available upon reasonable request to the authors

---

**Figure 1:**
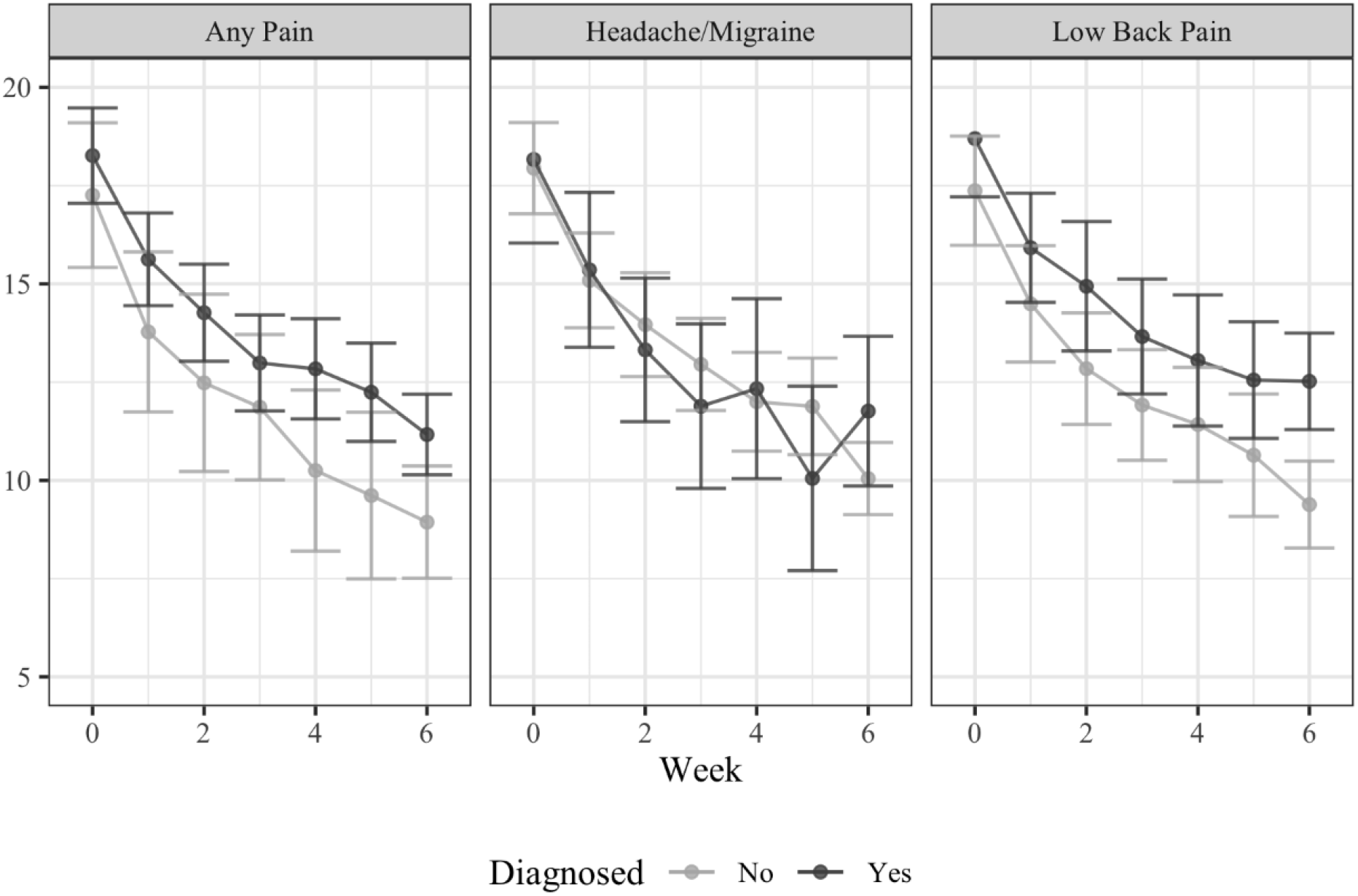
Change in depression symptomology (PHQ-9) among Veterans with and without a pain diagnosis.

**Supplemental Table 1:**
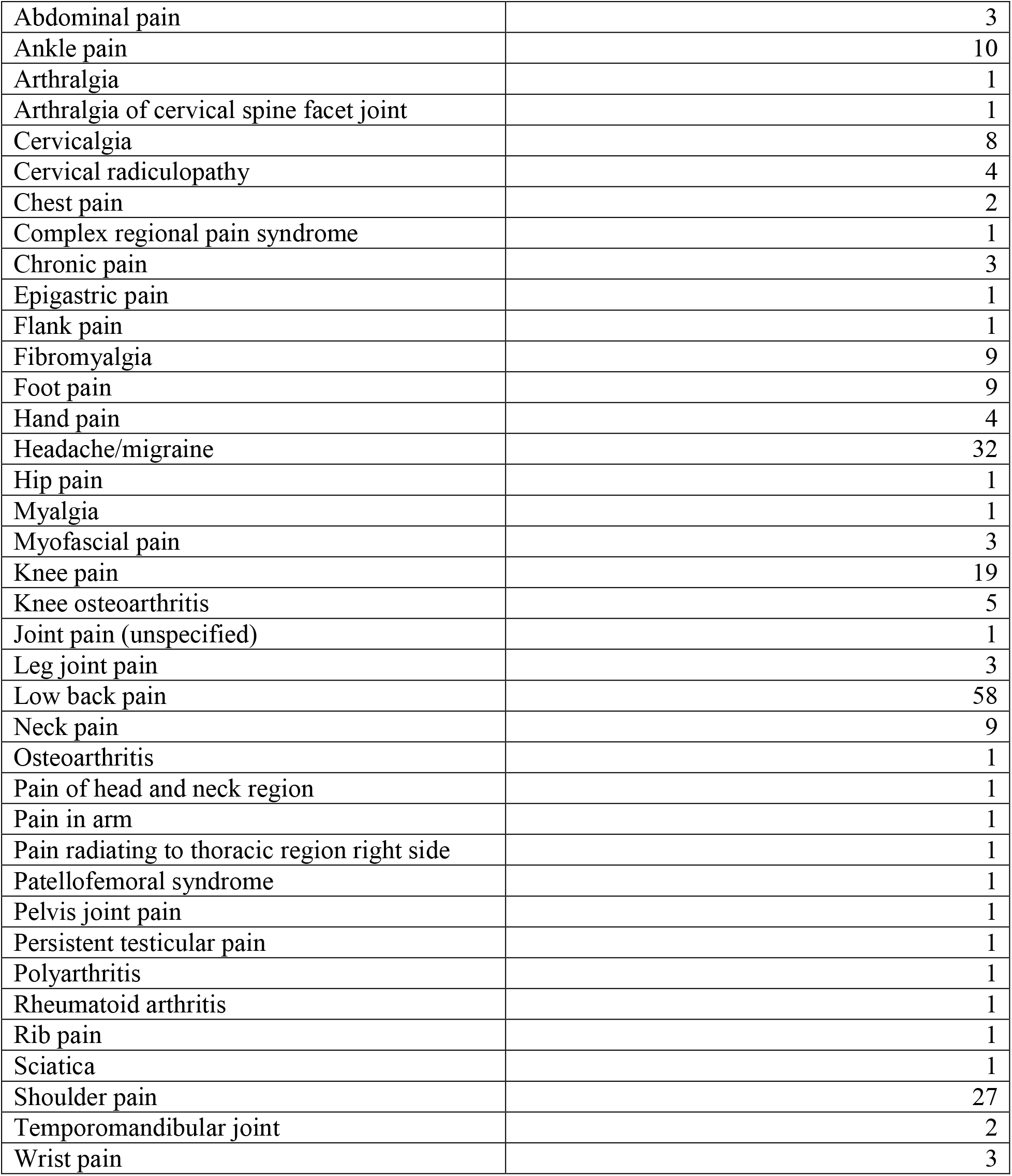
List of pain diagnoses from chart review.

