## supplemental table 1 for "Effects of Chronic Pain Diagnoses on the Antidepressant Efficacy of Transcranial Magnetic Stimulation"

| Abdominal pain | 3 |
| --- | --- |
| Ankle pain | 10 |
| Arthralgia | 1 |
| Arthralgia of cervical spine facet joint | 1 |
| Cervicalgia | 8 |
| Cervical radiculopathy | 4 |
| Chest pain | 2 |
| Complex regional pain syndrome | 1 |
| Chronic pain | 3 |
| Epigastric pain | 1 |
| Flank pain | 1 |
| Fibromyalgia | 9 |
| Foot pain | 9 |
| Hand pain | 4 |
| Headache/migraine | 32 |
| Hip pain | 1 |
| Myalgia | 1 |
| Myofascial pain | 3 |
| Knee pain | 19 |
| Knee osteoarthritis | 5 |
| Joint pain (unspecified) | 1 |
| Leg joint pain | 3 |
| Low back pain | 58 |
| Neck pain | 9 |
| Osteoarthritis | 1 |
| Pain of head and neck region | 1 |
| Pain in arm | 1 |
| Pain radiating to thoracic region right side | 1 |
| Patellofemoral syndrome | 1 |
| Pelvis joint pain | 1 |
| Persistent testicular pain | 1 |
| Polyarthritis | 1 |
| Rheumatoid arthritis | 1 |
| Rib pain | 1 |
| Sciatica | 1 |
| Shoulder pain | 27 |
| Temporomandibular joint | 2 |
| Wrist pain | 3 |

Supplemental Table 1: List of pain diagnoses from chart review.
